# Genetic risk score for ovarian cancer based on chromosomal-scale length variation

**DOI:** 10.1101/2020.07.18.20156976

**Authors:** Chris Toh, James P. Brody

**Affiliations:** Department of Biomedical Engineering, University of California, Irvine

## Abstract

**Introduction:** Twin studies indicate that a substantial fraction of ovarian cancers should be predictable from genetic testing. Genetic risk scores can stratify women into different classes of risk. Higher risk women can be treated or screened for ovarian cancer, which should reduce overall death rates due to ovarian cancer. However, current ovarian cancer genetic risk scores, based on SNPs, do not work that well. We developed a genetic risk score based on structural variation, quantified by variations in the length of chromosomes.

**Methods:** We evaluated this genetic risk score using data collected by The Cancer Genome Atlas. From this dataset, we synthesized a dataset of 414 women who had ovarian serous carcinoma and 4225 women who had no form of ovarian cancer. We characterized each woman by 22 numbers, representing the length of each chromosome in their germ line DNA. We used a gradient boosting machine, a machine learning algorithm, to build a classifier that can predict whether a woman had been diagnosed with ovarian cancer in this dataset.

**Results:** The genetic risk score based on chromosomal-scale length variation could stratify women such that the highest 20% had a 160x risk (95% confidence interval 50x-450x) compared to the lowest 20%. The genetic risk score we developed had an area under the curve of the receiver operating characteristic curve of 0.88 (estimated 95% confidence interval 0.86-0.91).

**Conclusion:** A genetic risk score based on chromosomal-scale length variation of germ line DNA provides an effective means of predicting whether or not a woman will develop ovarian cancer.

## Introduction

Ovarian cancer kills about 150,000 women per year worldwide[1]. The most common form of ovarian cancer, ovarian serous carcinoma is often diagnosed late (stage III (51%) or IV (29%)) and has a relatively bleak 5-year survival rate [2]. If women with an elevated risk of developing ovarian cancers could be identified, interventions could be taken that would reduce the number of women who die from ovarian cancer. These interventions include prophylactic oophorectomies, which would completely avoid ovarian cancer, and more targeted screening, which could identify ovarian cancers in earlier stages, where surgery is an effective cure[3–6]. These interventions could both increase 5-year survival times and reduce the overall number of deaths due to ovarian cancer.

A substantial fraction of ovarian cancers should be predictable by genetic testing. The heritability of ovarian cancer has been measured at about 40% (95% confidence interval 23%-55%) by the Nordic Twin Study[7]. The maximum discriminative accuracy of a genetic risk test is a function of both the heritability and the prevalence of the disease [8,9]. Based on the measured heritability (about 40%) and prevalence (about 0.1%) of ovarian cancer, the maximum accuracy, measured by the area under the receiver operating characteristic curve (AUC), should be greater than 0.95, where 1.0 indicates a perfect test. Current genetic risk scores do not approach that level of accuracy.

Most current genetic risk scores are derived from single nucleotide polymorphisms (SNPs) identified by genome wide association studies[10–15]. These tests, called polygenic risk scores, construct a score based on a linear combination of the value of a collection of SNPs. This strategy has been moderately successful with ovarian cancer. One study followed this strategy to construct a polygenic risk score where women who scored in the top 20% had a 3.4-fold increased risk compared to women who scored in the bottom 20%[16].

We developed an alternative strategy to compute genetic risk scores. Our strategy is based on structural variation rather than SNPs and uses machine learning algorithms, which include non-linear effects, rather than linear combinations.

## Methods

We tested this strategy with data from the Cancer Genome Atlas (TCGA) project. TCGA was a project sponsored by the National Cancer Institute to characterize the molecular differences in 33 different human cancers[17–19]. The project collected samples from about 11,000 different patients, all of whom were being treated for one of 33 different types of tumors. The samples collected usually included tissue samples of the tumor, tissue samples of normal tissue adjacent to the tumor and normal blood samples. (Normal blood samples were not available from patients diagnosed with leukemias.)

Most of the patient normal blood samples were processed to extract and characterize germline DNA. All germline DNA samples were processed by a single laboratory, the Biospecimen Core Resource at Nationwide Children’s Hospital. Single nucleotide polymorphisms (SNPs) were measured from the patient samples with an Affymetrix SNP 6.0 array. This SNP data was then processed (by the TCGA project) through a bioinformatics pipeline [20], which included the packages Birdsuite [21] and DNAcopy [22]. The result of this pipeline is, for each sample, a listing of a chromosomal region (characterized by the chromosome number, a starting location, and an ending location) and the associated value given as the “segmented mean value.” The segmented mean value is defined as the logarithm, base 2 of one-half the copy number. A normal diploid region with two copies will have a segmented mean value of zero.

NCI has provided most of the TCGA data on the Genomic Data Commons [23]. The copy number variation is called the masked copy number variation on the Genomic Data Commons. The masking process removes “Y chromosome and probe sets that were previously indicated to have frequent germline copy-number variation.” [20].

This research uses de-identified coded datasets produced by TCGA. Therefore it is not considered human subjects research.

We accessed the TCGA data through Google’s BigQuery, a cloud-based database. This resource is hosted and maintained by the Institute of Systems Biology [24]. We used the copy number segment (masked) table extracted from the Genomic Data Commons in February 2017. We also used information from the Biospecimen (extracted April 2017) and Clinical (extracted June 2018) tables. The copy number table contained all the information for the chromosome scale length variation data. The Biospecimen table was used to identify which samples were from normal blood (representing germ line DNA). The Clinical table provided information on the individual patient’s gender, race, and ovarian cancer status. Information in the different tables was tied together by the sample barcode parameter.

We used the statistical computer language R to query the BigQuery database, collect the data and manipulate it into different forms. We took extensive care to avoid typical problems that lead to falsely high AUCs in machine learning. For instance, we ensured that no data leakage occurred, which can lead deceivingly high AUCs when copies of a sample appear in both the training and test sets.

We used the H2O machine learning package in R to create machine learning models. H2O takes care of setting many of the proper default values, depending on whether the goal of the model is classification or regression. For the gradient boosting machine (GBM) models, H2O performs preprocessing, randomization, encoding categorical variables, and other data processing steps appropriate for the chosen model.

H2O has an automated machine learning algorithm, named AutoML[25]. Given a spreadsheet like-dataset, AutoML will run through four different machine learning algorithms and evaluate which provides the best models for the given problem. For each of the machine learning algorithms, it will evaluate several different hyperparameters. The process is limited by the amount of time devoted to it. After the allotted time, AutoML reports a scoreboard ranking the best algorithms. For the gradient boosting machine algorithm, we started with the default H2O settings. These default settings build trees to a maximum depth of five trees with a sample rate of 1 [26]. For the results reported in Table 2, we used an allotted time of one hour. In tests, we found that the results do not change substantially with times up to 10 hours.

We used 5-fold cross validation with the GBM algorithm to produce Table 3 and Figure 2. Cross validation uses repeated model runs with non-overlapping data. This approach allows one to use of all samples in the limited dataset. For Table 3 and Figure 2, we estimated 95% confidence intervals for the odds ratios following the method described in [27].

Figure 3 was produced with a single model run by splitting the dataset into a training set containing 80% of the data and a test set containing 20% of the data.

## Results

Using the TCGA dataset, we identified a measure that we call *chromosome-scale length variation*. Taken together, structural variations like insertions, deletions, translocations and copy number variations slightly alter the overall length of an individual’s chromosome. Thus, the lengths of the set of chromosomes can be used to characterize a person. A histogram showing the distribution of relative chromosome lengths taken from germ line DNA samples in the TCGA dataset is shown in Figure 1. By convention, these lengths are reported in units of log base 2. A value of “0” represents the consensus, average, chromosome length.

**Figure 1.**
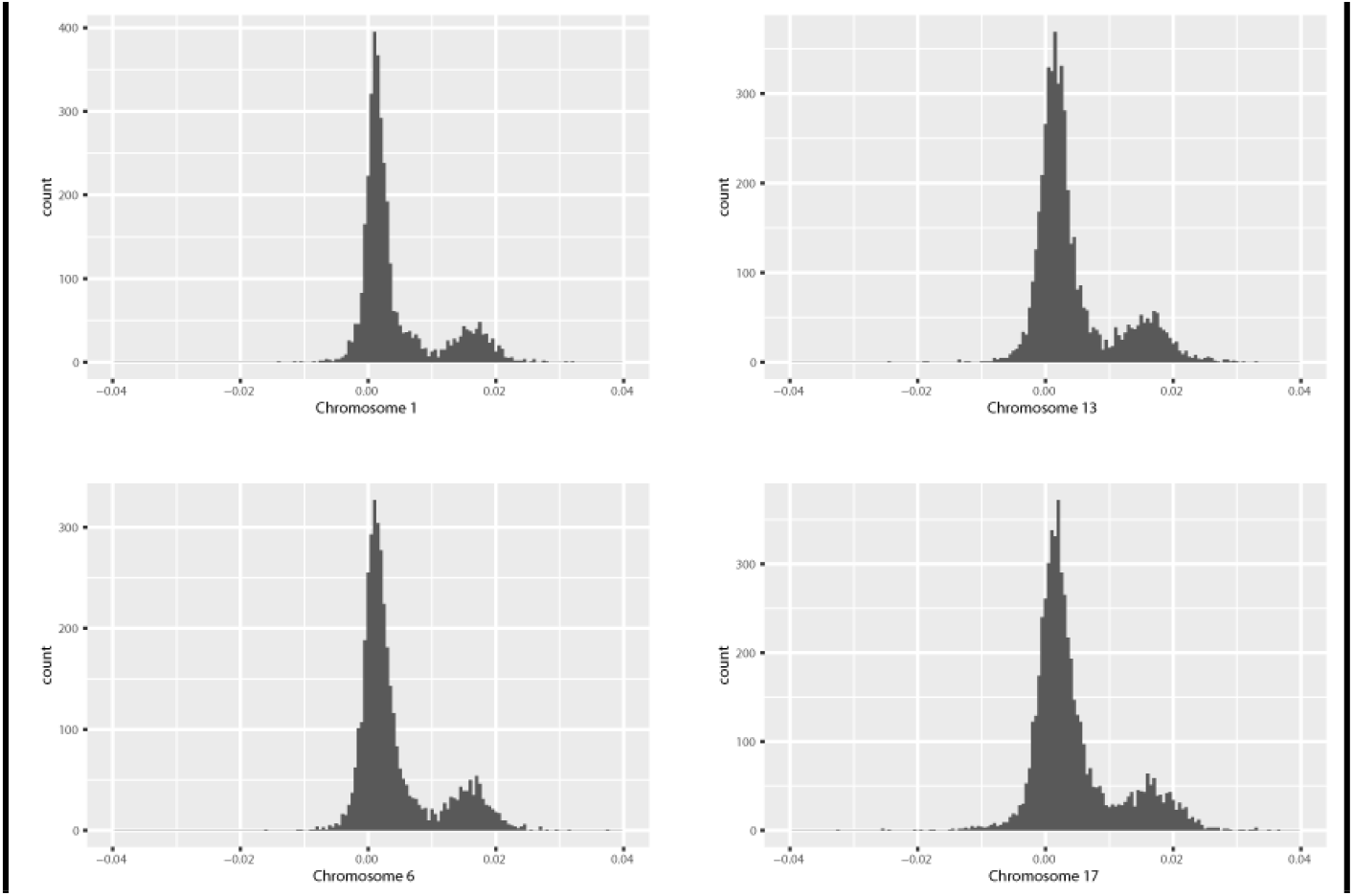
This shows a histogram of chromosome scale length variation for most of chromosome 17. For most patients in the TCGA dataset, a normal blood sample was taken, genomic DNA was extracted from that sample and analyzed with an Affymetrix SNP 6.0 array. The data from this array was processed by the TCGA project through a bioinformatic pipeline that resulted in a segment mean value, which is a number equal to the log base two of one half the copy number value. This histogram indicates that most people have a nominal value of 0, indicating exactly two copies of the diploid chromosome. A value of 0.02 indicates the person has on average 2.028 copies of the chromosome, or about 1.4% longer than the average length of the chromosome.

**Figure 2.**
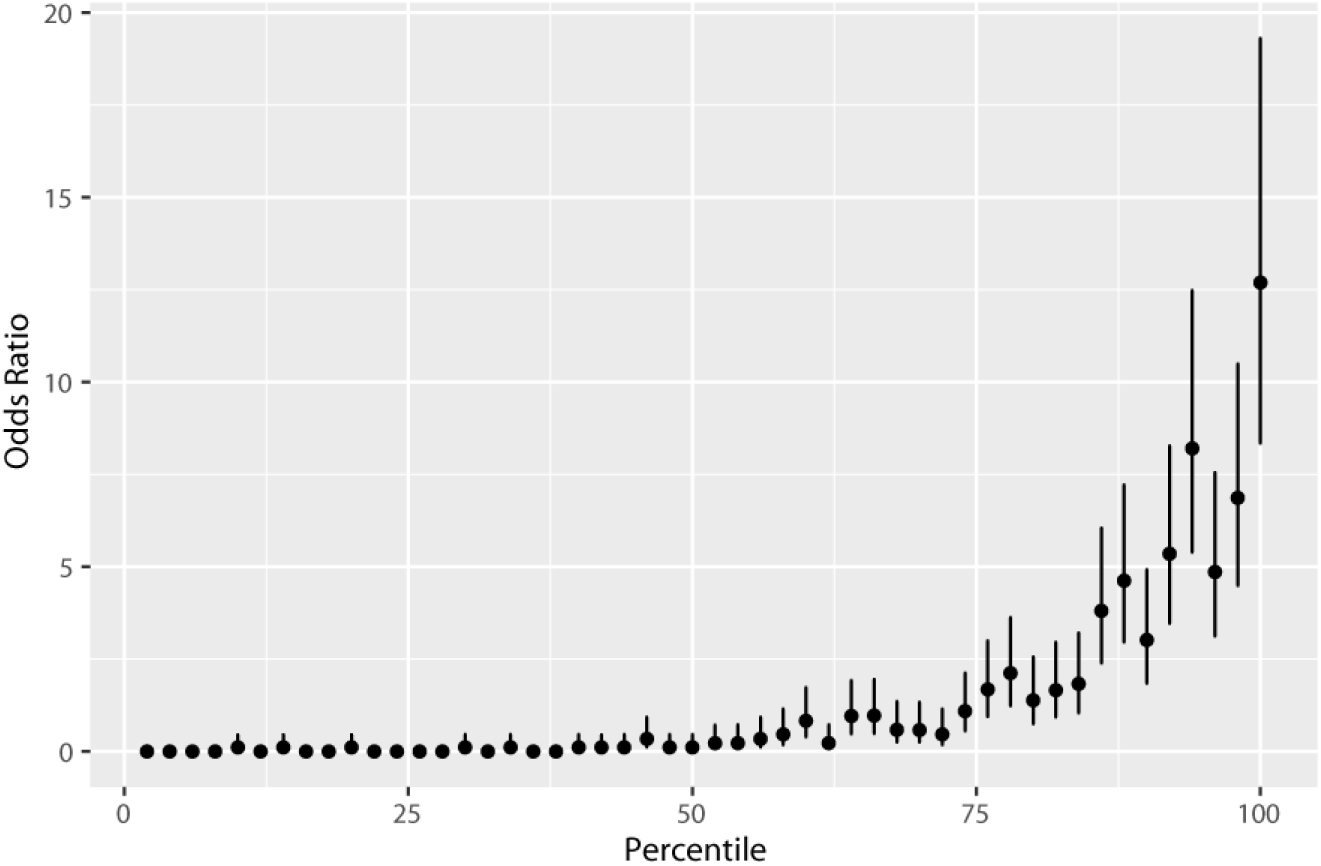
This figure shows that women ranked higher by the predictive model have significantly more likely to have ovarian cancer. The predictive model ranked all 4669 women in the dataset based on their likelihood of having ovarian cancer, based solely on germ line DNA data. This ranking was then split into 50 equal partitions, each with about 93 women. This plot shows the odds ratio (relative to 414 ovarian cases out of 4669 total) of each of the 50 equal partitions along with the 95% confidence intervals.

**Figure 3.**
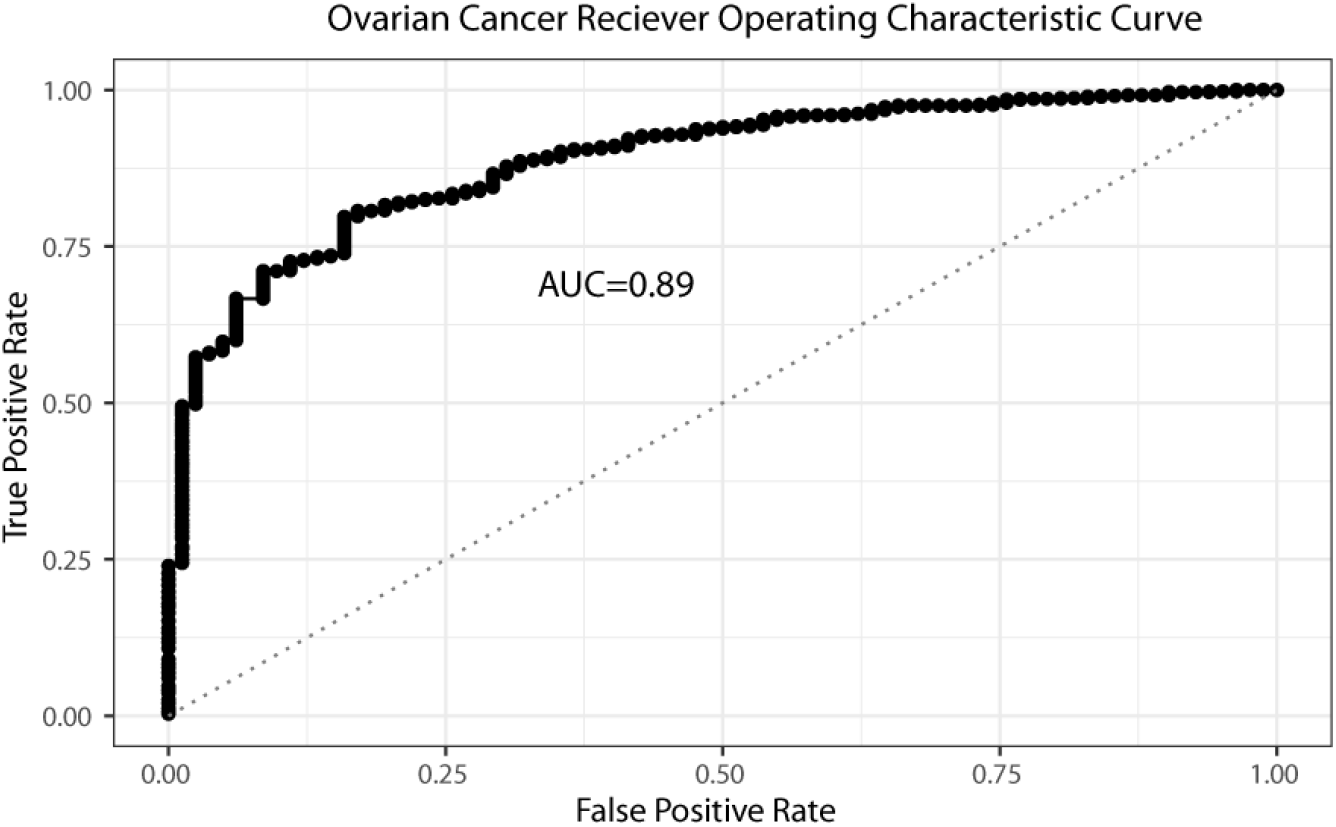
This figure presents a receiver operating characteristic curve of the model’s predictions. The area under the curve for this model was 0.89.

Figure 1. This figure shows a histogram of chromosome scale length variation for most of chromosomes 1,6,13, and 17. For most patients in the TCGA dataset, a normal blood sample was taken, genomic DNA was extracted from that sample and analyzed with an Affymetrix SNP 6.0 array. The data from this array was processed by the TCGA project through a bioinformatic pipeline that resulted in a segment mean value, which is a number equal to the log base two of one half the copy number value. This histogram indicates that most people have a nominal value of 0, indicating exactly two copies of the diploid chromosome. A value of 0.02 would indicate the person has on average 2.028 copies of the chromosome, or about 1.4% longer than the average length of the chromosome.

From the TCGA dataset, we synthesized a case-control study to test whether chromosome-scale length variation data can construct a genetic risk score. We identified 4225 women who had not been diagnosed with any form of ovarian cancer and 414 women who had been diagnosed with ovarian serous carcinoma. Statistical descriptions of the two populations are shown in Table 1.

**Table 1.**
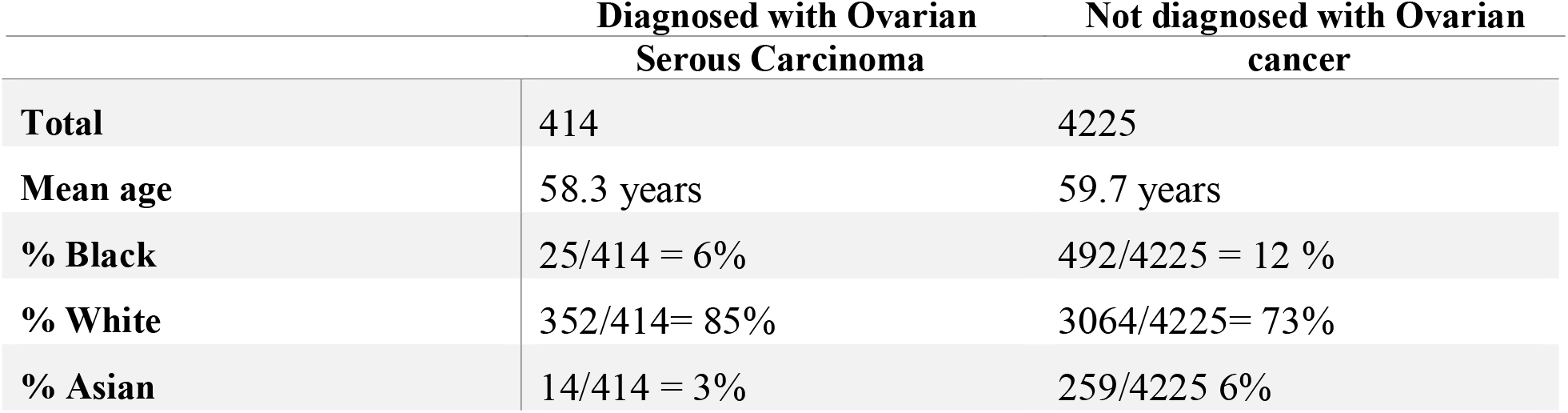
From the TCGA dataset, we constructed two groups, both solely composed of women. The first group, containing 414 women, all had been diagnosed with ovarian serous carcinoma. None of the second group, with 4225 women, had been diagnosed with any form of ovarian cancer. This table compares the two populations.

**Table 2.** This table lists five different machine learning algorithms we evaluated for predicting ovarian cancer from chromosome-scale length variation data using the H2O package in R. The algorithms are ranked by the best AUC it achieved using 5-fold cross validation.

| Algorithm | AUC |
| --- | --- |
| Gradient Boosting Machine | 0.88 |
| Distributed Random Forest | 0.87 |
| Extremely Randomized Trees | 0.86 |
| Deep learning | 0.82 |
| Generalized Linear Model | 0.68 |

**Table 3.** Using 5-fold cross validation, each woman in the dataset received a score from the model built to predict ovarian cancer. The women were ranked by score from lowest to highest and then partitioned into five quintiles. This table presents the number of women with and without ovarian cancer in each quintile along with the odds ratio (relative to the entire group) and the 95% confidence interval for the odds ratio.

| Quintile | Number of women without ovarian cancer | Number of women with ovarian cancer | Total number of women | Odds ratio | 95% confidence interval |
| --- | --- | --- | --- | --- | --- |
| 1 | 925 | 3 | 928 | 0.03 | 0.01--0.09 |
| 2 | 925 | 3 | 928 | 0.03 | 0.01--0.09 |
| 3 | 901 | 27 | 928 | 0.30 | 0.21--0.45 |
| 4 | 842 | 86 | 928 | 1.04 | 0.82--1.33 |
| 5 | 632 | 295 | 927 | 4.76 | 4.01--5.65 |

Next, we evaluated the effectiveness of several different machine learning algorithms. We measured how well these algorithms could classify a woman, based solely on the set of 23 chromosome-scale length variation measurements, into either the class with ovarian cancer or without. The measurement of success we used was the area under the curve (AUC) of the receiver operating characteristic curve. The results of these measurements are shown in Table 2.

Based on the results in Table 2, we used the Gradient Boosting Machine algorithm throughout the rest of this manuscript. In the next step, we sought to classify the 4669 women in the dataset. We used a *k*-fold cross validation procedure, with *k*=5. The dataset was randomly partitioned into five equal groups. The first group was held out (to be the test set), while the other four groups were used to train a model to distinguish the two classes (women with ovarian cancer and women without ovarian cancer). The trained model assigned a numerical score to each of the women in the first group (test set) quantifying how likely that woman was a member of the ovarian cancer class. The process was repeated 5 times, with a different group held out each time. The result is a numerical score for each of the 4669 women.

The predictions were compared to the known ovarian cancer status of each of the 4669 women. First, all 4669 women were ranked by their score, representing the likelihood that they were from the ovarian cancer class. By comparing this ranking with their known ovarian cancer status, we can evaluate how well the model classified the women.

The comparison is presented in two different forms. Table 3 provides a tabular form of relative risk for the population segmented into five different groups. Figure 2 shows similar information in graphical form, where the population is segmented into 50 groups.

Finally, we took the dataset of 4669 women and split it into a training set (80%) and a test set (20%). Using H2O, we trained a Gradient Boosting Machine model to predict whether a woman was in the group with ovarian cancer, or not. The results are presented in Figure 3, which shows a classic receiver operating characteristic curve of the model’s predictions.

Figure 2. This figure shows that women ranked higher by the predictive model are significantly more likely to have ovarian cancer. The predictive model ranked all 4669 women in the dataset based on their likelihood of having ovarian cancer, based solely on germ line DNA data. This ranking was then split into 50 equal partitions, each with about 93 women. This plot shows the odds ratio (relative to 414 ovarian cases out of 4669 total) of each of the 50 equal partitions along with the 95% confidence intervals.

Figure 3. This figure presents a receiver operating characteristic curve of the model’s predictions. The area under the curve for this model was 0.88.

## Discussion

The results presented here compare favorably to other genetic risk scores for ovarian cancer. For instance, a previous study found that a polygenic risk score in the top 20% conferred a 3.4-fold risk increase compared to women in the bottom 20% [16]. As seen in Table 3, the top 20% in our results had an increase of over 100-fold risk over women who scored in the bottom 20%.

Table 2 quantifies different algorithms applied to this problem. These results are illustrative, but not conclusive. Tuning machine learning models is an art, and it might be possible, for instance, to tune a deep learning network to obtain superior results. In similar work on TCGA colon cancer data, we found that a pairwise neuron network algorithm performs equal to a gradient boosting machine[28]. The gradient boosting machine generally runs faster and is easier to tune. Others have evaluated different machine learning algorithms for different bioinformatic problems and found that no one algorithm is superior[29]. They also found that a gradient boosting machine algorithm does perform well on many different types of datasets, consistent with out findings.

A disadvantage of this approach, compared to more conventional SNP-based genetic risk scores, is that the results are difficult to understand and extract biological meaning. The Gradient Boosting Machine computational model is complex, consisting of dozens of decisions trees. Furthermore, the data that is used to traverse the decision tree is also complex. The data consists of chromosome scale length variation, which is the result of many different insertions, deletions, translocations, and other structural changes. Polygenic risk scores based on SNPs are easy to interpret. One can identify how much each SNP contributes to the score and one can locate this SNP in the genome and understand the function of nearby genes that might change. Although this approach is lacking in explanatory power, its ultimate goal is predictive power.

We considered whether the results were due to two common problems faced by GWAS studies: batch effects or population stratification. We found it unlikely that our model is identifying batch effects rather than real effects. First, all samples were collected from the same tissue, blood. This eliminates one common source of batch effects, since the DNA extraction process is the same for each sample. Second, all samples were processed by the same laboratory, the Nationwide Children’s Hospital Biospecimen Core Resource, with the same type of instrument. This laboratory followed the same protocol throughout their processing phase. Finally, we looked up the batch history of each sample. The 424 ovarian cancer samples were processed in 15 separate batches. The non-ovarian samples were processed in several hundred different batches. For these reasons, we do not believe the results are due to batch effects.

Population stratification occurs in case/control studies when the cases and controls contain substantially different proportions of genetically discernable subclasses. Most TCGA samples were collected in the United States from a racially diverse group. For instance, over half the ovarian cancer samples were collected at five locations in the United States: Memorial Sloan Kettering, Washington University, University of Pittsburgh, Duke, and Mayo Clinic-Rochester. Table 1 lists demographic information about the two populations. Although the table does indicate slightly different proportions, by race, in the case and control groups, it does not seem to be different enough to account for the AUC observed.

This study has several weaknesses. First, the control population in this analysis is not randomly drawn from the general population, but instead consists of women who were part of the study because they were diagnosed with another form of cancer. Second, the results rely on a single dataset. The general applicability of this method would be better established if we were able to show that a model trained on one dataset would perform well on a second dataset that was collected independently. Demonstrating that a model is transferrable is a longer-term goal of ours.

Future work could refine this method to improve the predictive ability of this method. The AUC might be improved through several strategies, including feature engineering, for instance using sub-chromosomes rather than complete chromosomes, data augmentation strategies, and the inclusion of SNP data. Further work can also establish how robust the model is: can a model trained with the TCGA data be successfully applied to a person not in the TCGA dataset.

## Conclusion

A genetic risk score based on chromosomal-scale length variation of germ line DNA provides an effective means of predicting whether or not a woman will develop ovarian cancer. Several avenues are open to further improve the AUC of this genetic risk score test.

## Data Availability

The results published here are based upon data generated by the TCGA Research Network: http://cancergenome.nih.gov/.

## Competing Interests

None of the authors have any competing interests.

## Acknowledgements

The results published here are in whole or part based upon data generated by the TCGA Research Network: http://cancergenome.nih.gov/.

## Notes

### Competing Interest Statement

The authors have declared no competing interest.

### Funding Statement

No external funding has been received for this work.

### Author Declarations

This research involves analysis of de-identified data initially collected by The Cancer Genome Atlas Program. The UC Irvine Institutional Review Board reviewed this research and found that it is exempt since the research does not involve identifiable private information.

